# Association between prenatal alcohol exposure and children’s facial shape. A prospective population-based cohort study

**DOI:** 10.1101/2021.07.22.21260946

**Authors:** X. Liu, M. Kayser, S. A. Kushner, H. Tiemeier, F. Rivadeneira, V. W. V. Jaddoe, W. Niessen, E.B. Wolvius, G. V. Roshchupkin

**Affiliations:** Department of Radiology and Nuclear Medicine, Erasmus MC University Medical Center Rotterdam, Rotterdam, the Netherlands; Department of Oral and Maxillofacial Surgery, Erasmus MC University Medical Center Rotterdam, Rotterdam, the Netherlands; Department of Genetic Identification, Erasmus MC University Medical Center Rotterdam, Rotterdam the Netherlands; Department of Psychiatry, Erasmus MC University Medical Center Rotterdam, Rotterdam the Netherlands; Department of Social and Behavioral Science, Harvard T. H. Chan School of Public Health, Boston, MA, USA; Department of Internal Medicine, Erasmus MC University Medical Center, Rotterdam, the Netherlands; The Generation R Study, Erasmus MC University Medical Center Rotterdam, Rotterdam, the Netherlands; Department of Pediatrics, Erasmus MC University Medical Center Rotterdam, Rotterdam the Netherlands; Faculty of Applied Sciences, Delft University of Technology, Delft, the Netherlands; Department of Epidemiology, Erasmus MC University Medical Center Rotterdam, Rotterdam the Netherlands

## Abstract

**IMPORTANCE:** Children exposed to a high level of prenatal alcohol exposure (PAE) are more likely to develop fetal alcohol spectrum disorder with adverse phenotypes on their faces. However, it is still poorly understood, which level of PAE is associated with facial manifestation and if such associations persist during childhood.

**OBJECTIVE:** To examine the association between PAE and children’s facial phenotype in a prospective multi-ethnic population-based study.

**DESIGN, SETTING, AND PARTICIPANTS:** This study was based on the Generation R Study, a prospective cohort from fetal life onwards with maternal and offspring data. Children had a 3D facial image taken at ages 9 (n=3160) and 13 years (n=2492).

**EXPOSURES:** We defined 6 levels of PAE based on the frequency and dose of alcohol consumption, and defined three tiers based on the timing of alcohol exposure of the unborn child.

**MAIN OUTCOMES AND MEASURES:** For image analysis, we used 3D graph convolutional networks for non-linear dimensionality reduction, which compressed the high-dimensional images into 200 endophenotypes representing facial morphology. These facial endophenotypes were used as dependent variables in a linear regression analysis to search for associations with PAE. Finally, to detect specific facial components associated with PAE, we mapped statistically significant endophenotypes back to the 3D facial shape. We generated heatmaps to display the facial changes associated with PAE. PAE prediction based on facial shape was also performed; the prediction accuracy was estimated by area under the receiver operating characteristic curve (AUC).

**RESULTS:** A significant association between PAE and facial shape was found in the 9-year-old children, at all levels of alcohol exposure: the higher the level of exposure, the stronger the association. Moreover, PAE before and during pregnancy was associated with facial shape. The most common detected facial phenotypes included turned-up nose tip, shortened nose, turned-out jaw and turned-in lower-eyelid-related regions. For the 13-year-old children, the associations were weaker and the AUCs lower than those of the 9-years-old children.

**CONCLUSIONS AND RELEVANCE:** PAE before and during pregnancy, even at low level, is associated with the facial shape of children, and these associations become weaker as children grow older.

**Key points:** *Questions:* Which level of prenatal alcohol exposure (PAE) is associated with children’s facial shape at different lifetimes?

*Findings:* A 3D facial phenotype analysis of 9- (N=3160) and 13- (N=2492) year-old children was conducted. PAE before and during pregnancy, even at low level, associates with the children’s facial shape; the association becomes weaker as children grow older.

*Meaning:* The results suggest avoiding drinking alcohol at least three months before and during pregnancy is the safest option for the child. More research is needed to investigate the consequences of PAE on children; the discovered facial phenotypes could be used as a biomarker for further investigations.

## INTRODUCTION

High levels of prenatal alcohol exposure (PAE) during pregnancy can have significant adverse associations with a child’s health development resulting in fetal alcohol spectrum disorder (FASD). FASD is defined as a combination of growth retardation, neurological impairment and recognizably abnormal facial development.^1,2^ The association of low to moderate alcohol consumption with the child’s health is less known, but could still have severe consequences for the child’s health, including lower birth weight, smaller birth size and preterm birth.^3,44^

Facial morphology can serve as a biomarker for health conditions and indicate developmental problems.^4, 5, 28, 29^ Since low to moderate alcohol consumption during pregnancy is associated with the child’s development, it might be also associated with the facial morphology. However, the results of previous research are ambiguous.

Douglas^6^ summarized the main approaches to detect the association between PAE and the human face, and concluded that compared with direct craniofacial anthropometry^7^ or measurement from photographs, ^8^ 3D surface imaging is the most promising way to reduce measurement error. For this reason, Muggli et al.^9^ performed a point-to-point 3D face registration analysis in a moderately sized study of 434 on 12-month-old infants. After adjusting for potential covariates, significant facial trait associations were found at low to moderate levels of PAE, mainly with shape of the jaw, forehead, nose, and areas near eyes. Howe et al.^10^ performed a landmark-based distance measurement on the 3D face in 4233 children (mean age: 15.4), but found no evidence of association with PAE. One major limitation of the landmarking approach used by Howe et al. is that it does not capture the complexity of facial morphology. Additionally, the participants in the Howe et al. study were much older (mean age: 15.4) than those in the study of Muggli et al. and association of PAE with the facial morphology might attenuate during childhood and adolescence.^17,18^ The present study addresses these limitations.

Recently, deep neural networks (DNNs),^11^ a data-driven method, which can extract key information from high-dimensional input data, has become the state-of-the-art method for clinical applications^12–14^. One type of DNN architecture used for dimensionality reduction is the auto-encoder,^15^ typically consisting of an encoder and decoder. The encoder is able to compress the high-dimensional 3D facial shape into low-dimensional representations of the facial morphology. The decoder then resamples these representations back to reconstruct the 3D facial shape.

In this paper, we applied a deep learning algorithm to 3D photographs of children from a multi-ethnic prospective pregnancy cohort. We used an auto-encoder to reduce the facial complexity and then examined association of low-dimensional representation with PAE before and during pregnancy. Furthermore, we also attempted to predict PAE from children’s facial shape, in order to test if facial morphology can be a biomarker that provides additional and independent clues for PAE diagnosis.

## METHODS

### Design and study population

This study was embedded in the Generation R Study. This is an ongoing population-based cohort study of pregnant women and their children from fetal life onwards. The goal is to identify early environmental and genetic causes leading to normal and abnormal growth, development, and health.^16^ All women living in the study area of Rotterdam, the Netherlands, who delivered between April 2002 and January 2006 were eligible. The Medical Ethical Committee approved the study and all participants gave written informed consent. A total of 9,778 participants were enrolled in the Generation R Study. 3D facial images of the children were taken at 9 and 13 years old, 5423 and 4551 respectively. The population consisted of 17 different ethnicities. We selected 4 major ethnicities (Dutch, non-Dutch Western, Turkish and Moroccan) and clustered them into 2 groups: Western (including Dutch and non-Dutch Western) and non-Western (including Turkish and Moroccan). After data cleaning, 3160 9-year-old children and 2492 13-year-old children were included in our study, with 1878 children assessed at both ages.

### Alcohol measurements

Information about maternal alcohol consumption was obtained by postal questionnaires in early, mid-, and late pregnancy.^3^ Mothers who reported any drinking were asked to classify their average alcohol consumption into one of the following 6 levels: <1 drink per week; 1–3 per week; 4–6 per week; 1 per day; 2–3 per day; >3 per day. An average alcoholic drink contains about 12 g of alcohol.

We defined 3 tiers to understand the association of alcohol exposure in different pregnancy stages. In all tiers, mothers who were abstinent before and during pregnancy comprised the control group. Settings of the exposed groups are shown in **Figure 1**. Tier 1 defined mothers only drinking 3 months before pregnancy as the exposed group, while tier 2 defined mothers drinking during pregnancy as the exposed group. In Tier 2a, mothers who drank during the first trimester of pregnancy, but were abstinent during the rest trimesters constituted the exposure group. Tier 2b followed similar exposure definitions as Tier 2a, but included mothers who also drink during the rest trimesters in the exposed. It is worth noting that 99% of mothers who drank during pregnancy also drank 3 months before pregnancy.

**Figure 1:**
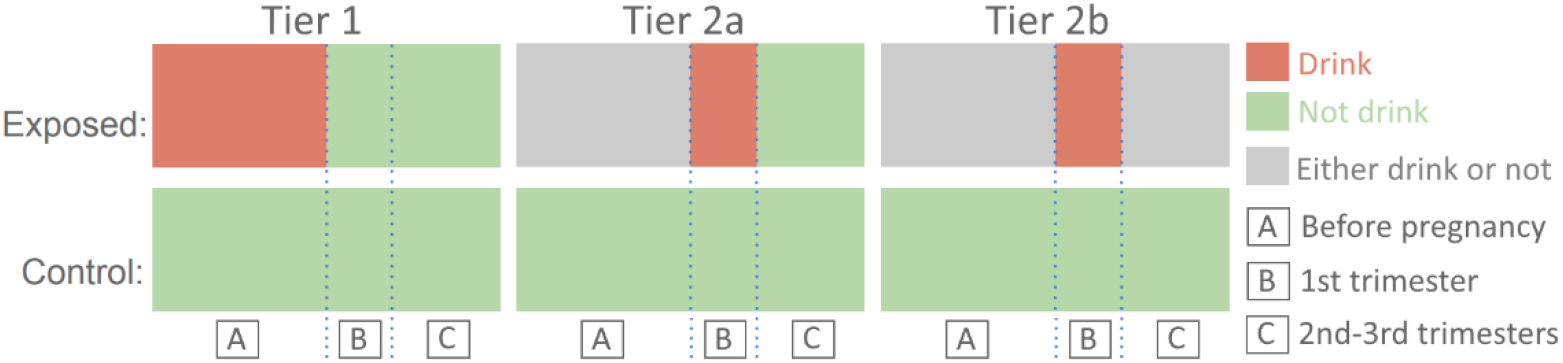
Definition of Tier 1, Tier 2a and Tier 2b.

### Data preprocessing

The 3D face images were collected with the 3dMD cameras system (3dMD Corp). The Distance and angle between the participants and cameras were fixed when taking photos. We adopted 3D morphology registration pipelines^45^ to build the raw data into a template-based dataset, in which each facial shape was modelled by a 3D Graph^27^ with the same vertex number and edge connectivity. More details about the data preprocessing can be found in the **Supplementary materials**.

### Dimensionality reduction to generate facial endophenotypes

We used a 3D graph auto-encoder^21^ for high-dimensional facial shape analysis. As shown in **Figure 2**, the auto-encoder consists of Encoder and Decoder that can perform feature mapping in a non-linear manner. The Encoder compresses the high-dimensional 3D facial shape into N latent features, while the Decoder reconstructs the 3D facial shape from the latent features. By minimizing the error between input and reconstructed facial shapes, the main morphology of the facial shape is captured in the N latent features. The encoding and decoding process can be formulated as:

**Figure 2:**
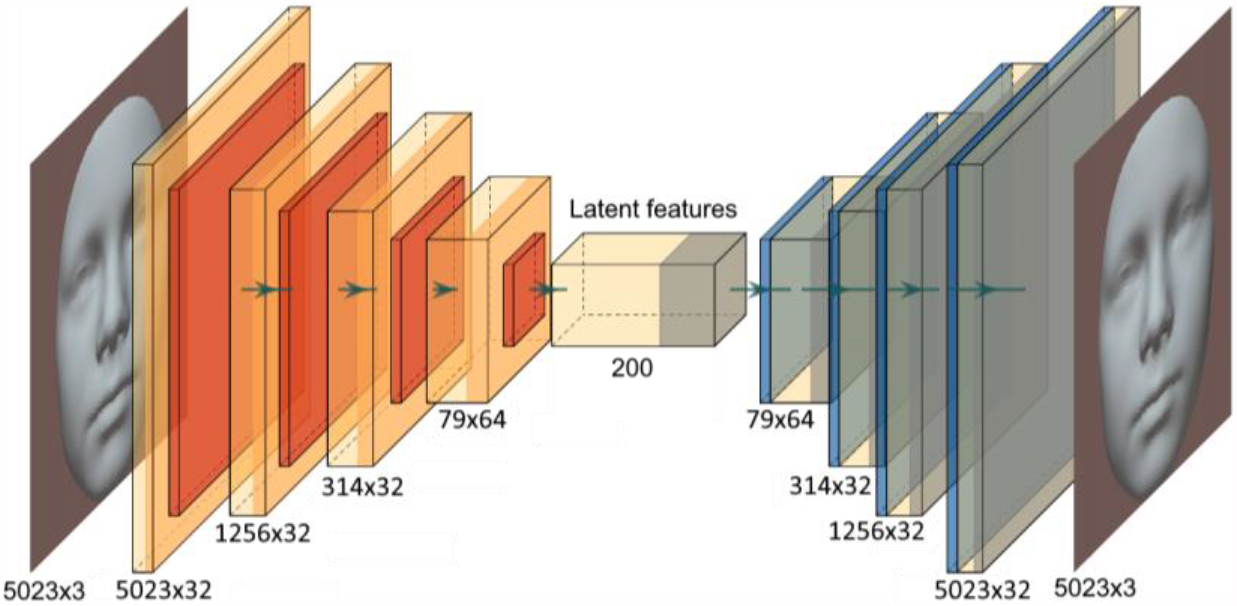
Framework of the auto-encoder. Red: down-sample process; Blue: up-sample process.

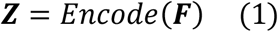

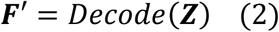

where ***Z***=[*Z*_0_, *Z*_1_, …, *Z*_N_] refers to the N latent features, *Encode*() and *Decode*() refer to the down-sampling and up-sampling process respectively. ***F*** denotes the input 3D facial shape, while ***F***′ represents the reconstructed 3D facial shape. In this paper, we defined these N latent features as N facial endophenotypes. In order to make the trade-off between reconstruction error and dimensional complexity, we conducted experiments on different numbers of endophenotypes. The optimum number was found to be 200. As shown in **Figure S3** (**Supplementary**), each endophenotype represents specific facial phenotypes. A uniform measurement *f*(*Z*) was defined in **Equation S1** (**Supplementary**), to measure the effect size of each endophenotype on the facial shape. More implementation details can be found in the **Supplementary materials**.

### Statistical analysis

After the dimensionality reduction each 3D facial image is represented by 200 endophenotypes. We performed linear regression analysis where each endophenotype was entered as dependent variable. The regression covariates included potential confounders: ethnicity, maternal age, maternal smoking in pregnancy, children BMI, age and gender.

We ran independent linear regression models for 200 facial endophenotypes. In order to correct the p-value for multiple testing, we calculated the false discovery rate (FDR) with **α =** 0.05. We selected FDR-significant endophenotypes and mapped them back to the 3D facial shape. **Figure S3** & **S4** further explain details about the mapping and visualization of selected endophenotypes on the facial shape.

The analysis was performed to examine four factors: tier of exposure, level of exposure, child ethnicity, and age. We first studied different tiers with PAE level in the exposed >1 (**Figure 5**), and compared different levels of exposure for Tier 2b (**Figure 4** & **S6**). Then all other results were based on Tier 2b within PAE level in the exposed >1. For the ethnicity factor, we defined 2 groups based on questionnaire information: the multi-ethnic group (including Dutch, non-Dutch Western, Turkish and Moroccan) and the Western group (including Dutch and non-Dutch Western background). We performed the analysis separately in the 9-year-old children, the 13-year-old children, and defined ‘growth’. The ‘growth’ was defined as the endophenotype differences between the 9- and 13-year-old children, including children assessed twice. The endophenotypes of ‘growth’ were computed by:

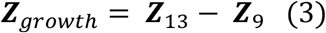

where ***Z***_*growth*_ refers to the growth of endophenotypes, ***Z***_13_ and ***Z***_9_ are endophenotypes of 13-year-old and 9-year-old children, respectively.

### Phenotypes recognition for PAE

The PAE prediction was only performed in children of Dutch national origin. We used logistic regression to model binary prediction, where 200 facial endophenotypes were used for prediction of PAE. Non-exposed children were set as the control group, while children with PAE level > 1 were set as exposed group. The prediction accuracy was quantified with the area under the receiver operating characteristic curve (AUC), with 5-fold cross validation. In order to adjust for potential biases, we firstly set the above-mentioned covariates as independent variables and obtained the baseline results (model A). Then we added facial endophenotypes as independent variables to the baseline model (model B).

*F*-test is often used to identify the model that best fits the population from which the data were sampled.^30^ Here we performed the analysis of variance (ANOVA) *F*-test on model A and model B, and then determined the F-value as well as p-value for each independent variable.

## RESULTS

The characteristics of the study population are summarized in **Table 1**. Maternal smoking, maternal age, child BMI and especially ethnicity showed imbalanced distribution between the control and exposed groups.

**Table 1:** Characteristics of children and their mothers included in the analysis.

**(a) 9-year-old children.**
|  |  | PAE Tier 1 | PAE Tier 2a | PAE Tier 2b |
| --- | --- | --- | --- | --- |
| characteristic | Control (Abstinent)<br>(N=760) | Exposed<br>(N=706) | Exposed<br>(N=1008) | Exposed<br>(N=1683) |
| Ethnicity (%) |  |  |  |  |
| Western |  |  |  |  |
| Dutch | 328(43.2) | 608(86.1) | 872(86.5) | 1476(87.7) |
| Non-Dutch Western | 39(5.1) | 74(10.5) | 119(11.8) | 181(10.8) |
| Non-Western |  |  |  |  |
| Turkish | 212(27.9) | 20(2.8) | 7(0.7) | 16(1.0) |
| Moroccan | 181(23.8) | 4(0.6) | 10(1.0) | 10(0.6) |
| Child's gender, No. (%) |  |  |  |  |
| Male | 357(47.0) | 306(43.3) | 489(48.5) | 823(48.9) |
| Female | 403(53.0) | 400(56.7) | 519(51.5) | 860(51.1) |
| Child BMI, mean (SD) | 18.6(3.2) | 17.4(2.6) | 17.1(2.2) | 17.0(2.1) |
| Child age, mean (SD), year | 9.8(0.4) | 9.7(0.3) | 9.8(0.3) | 9.8(0.3) |
| Maternal smoking, No. (%) |  |  |  |  |
| No | 556(73.2) | 406(57.5) | 508(50.4) | 846(50.3) |
| Yes | 204(26.8) | 300(42.5) | 500(49.6) | 837(49.7) |
| Maternal age, mean (SD) | 28.2(5.0) | 30.4(4.7) | 31.2(4.3) | 31.7(4.1) |

**(b) 13-year-old children.**
|  |  | PAE Tier 1 | PAE Tier 2a | PAE Tier 2b |
| --- | --- | --- | --- | --- |
| characteristic | Control (Abstinent)<br>(N=519) | Exposed<br>(N=579) | Exposed<br>(N=822) | Exposed<br>(N=1379) |
| Ethnicity (%) |  |  |  |  |
| Western |  |  |  |  |
| Dutch | 237(45.7) | 502(86.7) | 714(86.9) | 1206(87.5) |
| Non-Dutch Western | 33 (6.4) | 61(10.5) | 95(11.6) | 152(11.0) |
| Non-Western |  |  |  |  |
| Turkish | 122(23.5) | 11(1.9) | 10(1.2) | 17(1.2) |
| Moroccan | 127(24.5) | 5(0.9) | 3(0.4) | 4(0.3) |
| Child's gender, No. (%) |  |  |  |  |
| Male | 247(47.6) | 263(45.4) | 407(49.5) | 690 (50.0) |
| Female | 272(52.4) | 316(54.6) | 415(50.5) | 689 (50.0) |
| Child BMI, mean (SD) | 21.1(4.0) | 19.6(3.3) | 19.1(2.7) | 19.0(2.6) |
| Child age, mean (SD), year | 13.7(0.4) | 13.55(0.3) | 13.6(0.3) | 13.6(0.3) |
| Maternal smoking, No. (%) |  |  |  |  |
| No | 372(71.7) | 346(59.8) | 417(50.7) | 708 (51.3) |
| Yes | 147(28.3) | 233(40.2) | 405(49.3) | 671 (48.7) |
| Maternal age, mean (SD) | 28.2(5.1) | 30.7(4.7) | 31.2(4.2) | 31.7 (4.1) |

The results of the linear regression in the 9-year-old children survived correction for multiple testing with FDR. PAE before pregnancy (Tier 1) and during pregnancy (Tier 2a & 2b) was associated with facial endophenotypes (**Table 2**). **Table S2** further shows results of dose-response assessment for different levels of PAE. No FDR-significant results were found in the 13-year-old children, or for the ‘growth’ in the longitudinal analysis. The nominal significant results (p-value < 0.05) for 9-year-old, 13-year-old and ‘growth’ can be found in **Table S3**.

**Table 2:** Details about FDR-significant endophenotypes in the multi-ethnic group. 9-year-old children for PAE level > 1. Ne refers to the number of the exposed samples, while Nc refers to the number of the control samples. As defined in Equation S1 (Supplementary), f(z) is the effect size of the endophenotype on the facial shape. Facial endophenotype index with ‘W’ means they are also significant in only Western group.

| Endophenotype Index | p-value | FDR-corrected p-value | coefficient | SE | Mean | Std | f(z) |
| --- | --- | --- | --- | --- | --- | --- | --- |
| 29 (W) | 8.3e-08 | 1.7e-05 | 0.021 | 0.0040 | 0.008 | 0.052 | 197.95 |
| 44 (W) | 1.8e-04 | 0.020 | -0.014 | 0.0038 | 0.007 | 0.048 | 165.52 |
| 173 | 4.2e-04 | 0.030 | -0.014 | 0.0039 | -0.006 | 0.050 | 185.23 |

**(b)** Tier 2a (any PAE during first trimester, but abstinent during the rest trimesters). Ne = 563, Nc = 760
| Endophenotype Index | p-value | FDR-corrected p-value | coefficient | SE | Mean | Std | f(z) |
| --- | --- | --- | --- | --- | --- | --- | --- |
| 36 (W) | 4.2e-06 | 8.5e-04 | 0.021 | 0.0046 | 0.010 | 0.064 | 269.78 |
| 29 | 3.8e-05 | 0.0038 | 0.014 | 0.0035 | 0.008 | 0.052 | 197.95 |
| 139 (W) | 1.0e-04 | 0.0069 | -0.014 | 0.0035 | 0.007 | 0.048 | 189.49 |
| 173 | 2.8e-04 | 0.0138 | -0.013 | 0.0035 | -0.006 | 0.050 | 185.23 |
| 51 | 5.7e-04 | 0.0230 | -0.030 | 0.0087 | -0.002 | 0.125 | 833.34 |
| 87 (W) | 6.2e-04 | 0.0206 | -0.011 | 0.0032 | 3.5e-04 | 0.044 | 155.85 |
| 69 | 9.5e-04 | 0.0272 | 0.019 | 0.0059 | -0.017 | 0.080 | 485.34 |
| 125 | 1.5e-03 | 0.0381 | 0.015 | 0.0047 | 0.016 | 0.066 | 287.89 |

**(c)** Tier 2b (any PAE during first trimester). Ne = 756, Nc = 760
| Endophenotype Index | p-value | FDR-corrected p-value | coefficient | SE | Mean | Std | f(z) |
| --- | --- | --- | --- | --- | --- | --- | --- |
| 36 (W) | 7.1e-05 | 0.014 | 0.017 | 0.0044 | 0.010 | 0.064 | 269.78 |
| 139(W) | 9.3e-05 | 0.009 | -0.013 | 0.0033 | 0.007 | 0.048 | 189.49 |
| 29 | 1.9e-04 | 0.013 | 0.013 | 0.0034 | 0.008 | 0.052 | 197.95 |
| 51 | 2.5e-04 | 0.012 | -0.030 | 0.0083 | -0.002 | 0.125 | 833.34 |
| 69 | 5.6e-04 | 0.023 | 0.019 | 0.0055 | -0.017 | 0.080 | 485.34 |
| 173 | 8.9e-04 | 0.030 | -0.011 | 0.0033 | -0.006 | 0.050 | 185.23 |
| 87 (W) | 1.3e-03 | 0.036 | -0.010 | 0.0031 | 3.5e-04 | 0.044 | 155.85 |
| 57(W) | 1.9e-03 | 0.048 | 0.022 | 0.0070 | -0.013 | 0.101 | 648.42 |

Each endophenotype (index 0-199) represents specific facial phenotypes, and the facial interpretation of each single endophenotype can be found in **Figure S3**.

### Visualization of results

**Figure 3 & 4** respectively show the results of different ethnic groups, and results of different PAE levels, that survive multiple testing correction. **Figure 5** & **S5** shows the nominal significant results (p-value < 0.05) of different tiers, and of different ages (9-year-old, 13-year-old and the ‘growth’), respectively. For all heatmaps, red areas refer to inner changes while blue areas refer to outer changes towards the geometric center of the facial shape. **Figure S3** & **S4** explain how each heatmap was generated, by combing represented phenotypes of each significant endophenotype using their coefficients as weights. The most common detected facial phenotypes included turned-up nose tip, shortened nose, turned-out jaw and turned-in lower-eyelid-related regions.

**Figure 3:**
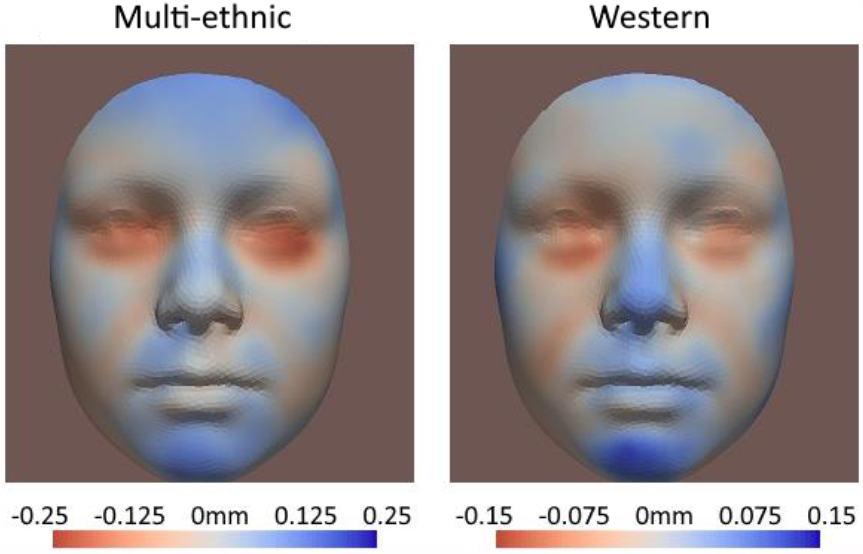
FDR-significant results of the multi-ethnic (Dutch, non-Dutch Western, Turkish and Moroccan) and the Western groups (Dutch and non-Dutch Western). Tier 2b (any PAE during first trimester), PAE level > 1 for 9-year-old children.

**Figure 4:**
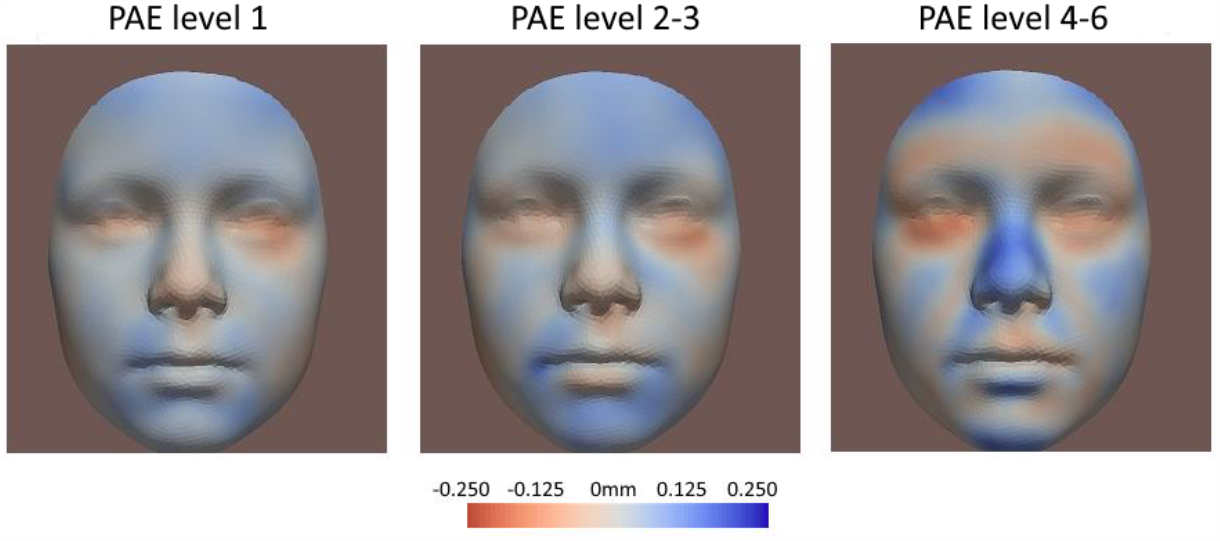
FDR-significant results of different levels of PAE. Tier 2b, multi-ethnic group for 9-year-old children. Level 1: <12 g of alcohol per week, N=887; Level 2-3: 12-72 g per week, N=643; Level 4-6: >12 g per day, N=79. Mothers who ever drank heavily (> 72 g a day) were excluded.

**Figure 5:**
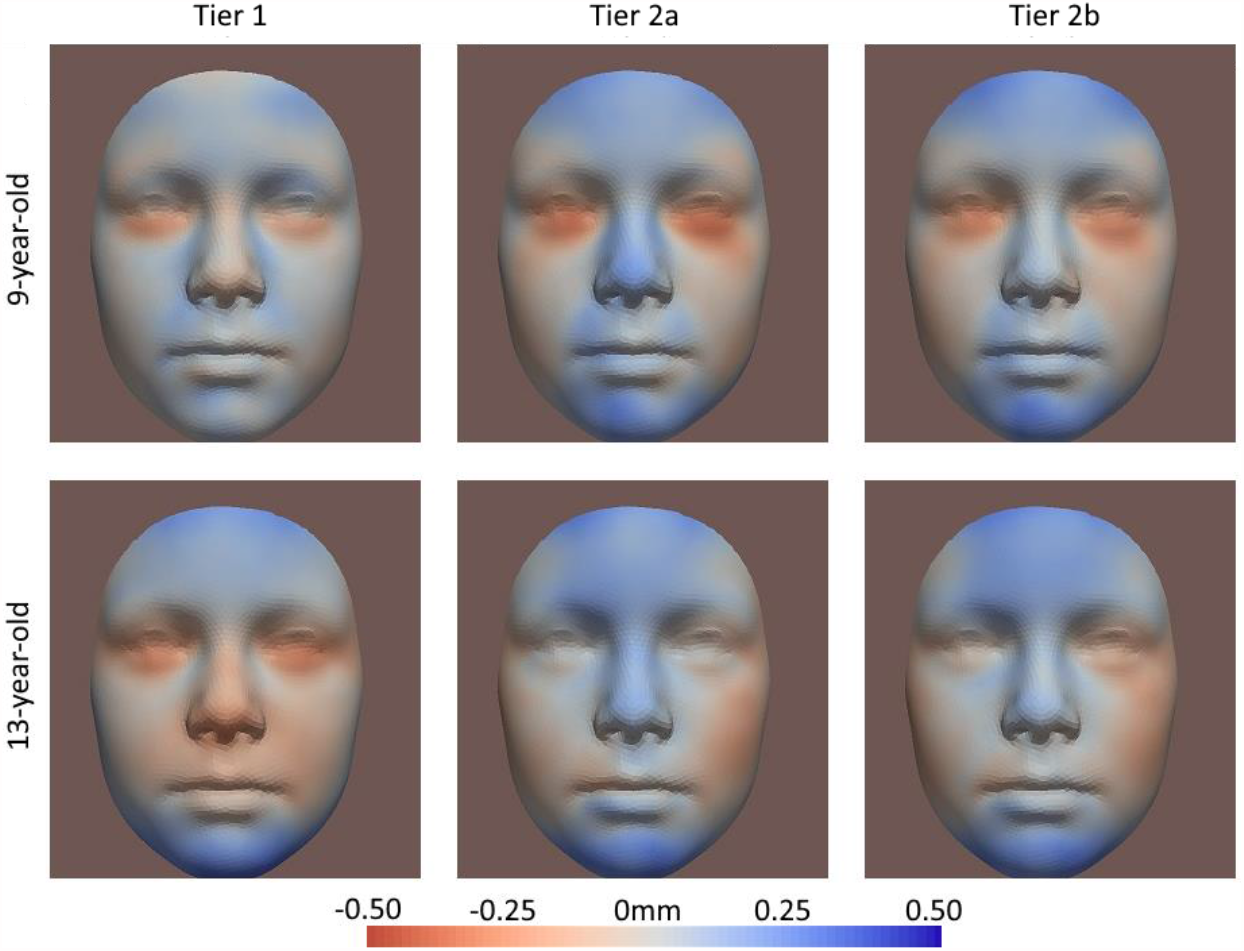
Nominal significant results of different tiers. Multi-ethnic, PAE level > 1. Tier 1: PAE only before pregnancy; Tier 2a: any PAE during first trimester, but abstinent during the rest trimesters; Tier 2b: any PAE during first trimester.

### Phenotypes recognition for PAE

**Table 3** shows the prediction results of the logistic regression, where model B with facial endophenotypes as independent variables obtained slightly higher AUC than model A (baseline). **Table S4** further shows the odds rations (OR) of endophenotypes in the logistic regression. The highest OR is 1.25 (P=.008; endophenotype index *36*) and 1.36 (P=.004; endophenotype index *14*) for 9- and 13-year-old children, respectively. **Table S5** shows the ANOVA *F*-test results, which confirmed the prediction model was improved when facial endophenotypes were included.

## DISCUSSION

This study examined the association between PAE and children’s facial shape by performing a multi-ethic population-based analysis, using state-of-the-art image analysis methodology including deep-learning approaches. A significant association between fetal alcohol exposure and facial morphology was found in the 9-year-old children, with a dose-response relationship (**Figure 4** & **S6, Table S2**): higher levels of alcohol exposure showed stronger associations. The most common detected facial phenotypes included turned-up nose tip, shortened nose, turned-out jaw and turned-in lower-eyelid-related regions.

**Table 3:** PAE prediction AUC with 5-fold cross validation. For children of Dutch national origin, PAE level > 1, Tier 2b. Ne refers to the number of the exposed samples, while Nc refers to the number of the control samples.

| Models | Independent variables | AUC in 9-year-old | AUC in 13-year-old |
| --- | --- | --- | --- |
|  |  | (Ne = 670, Nc = 329) | (Ne = 543, Nc = 236) |
| Model A | maternal age, maternal smoking in pregnancy, children BMI, age and gender | 0.769 | 0.756 |
| Model B | all independent variable in Model A, and 200 facial endophenotypes | 0.782 | 0.757 |

The association between low levels of PAE and children’s facial shape has been previously reported, but our study found an association at a much lower dose of exposure. Muggli et al.^9^ found a significant association at a low PAE level < 70 g of alcohol per week, in 12-month-old babies. Iveli et al.^43^ considered a light consumption < 700 mL per trimester (roughly < 46 g per week) and found that 66% of 79 newborns in the exposed group had some facial abnormality. However, in Tier 2b which assessed the dose-response (**Figure 4** & **Table S2**), where mothers who ever drank heavily (> 72 g a day) were excluded, we found that even if mothers drank very little (< 12 g per week) during pregnancy, the association between PAE and children’s facial shape could be observed.

The associations of PAE with children’s facial shape attenuated as children became older. As the results of different ages shown in **Figure S5**, for some phenotypes, the association we observed in 9-year-old children, were attenuated in the 13-year-old children. These changes were consistent with the heatmaps of ‘growth’, where ‘growth’ was defined endophenotype differences between the 9- and 13-year-old children (**Equation 3**). Besides, the results of Muggli et al. and ours differ from those of Howe et al.,^10^ where no association was found in the 15-year-old children. Possibly this discrepancy can be explained by a further attenuation of the association. Our results suggest that as children grow up, the association of PAE with children’s facial shape become weaker. This finding is consistent with the associations of PAE with children’s weight, height, and head circumference that attenuate as children become older.^17,18^ One possible explanation for this change over lifetime might be the impact of the environment. With age some alcohol-related phenotypes on the facial shape of children might be obscured by environmental influences, especially at the peak age of facial development (12-14 years for boys and 10-12 years for girls).^35,36^ Further investigations on the mechanism of association are needed to fully understand how the association between PAE and children’s facial shape develop and then attenuated with age.

The associations of PAE with children’s facial shape were similar in Tier 2a & 2b (Tier 2a: any PAE during first trimester, but abstinent during the rest trimesters; Tier 2b: any PAE during first trimester). This suggests that the associations were mainly explained by alcohol exposure during the first trimester of the pregnancy. Besides, we also examined the association of PAE before pregnancy with children’s facial shape. A significant association was found in Tier 1 (PAE only before pregnancy), but weaker than those of Tier 2 (PAE during pregnancy). It is worth noting that 99% of drinking mothers in Tier 2 also drank before pregnancy, but none of those drinking in Tier 1 also drank during pregnancy. Thus, the comparison (**Table 2** & **Figure 5**) between Tier 1 and Tier 2 indicated that associations of PAE with children’s facial shape were stronger in mothers who continued to drink during pregnancy (Tier 2), than in those who stopped when becoming pregnant (Tier 1). To the best of our knowledge, this study is the first to examine the association between PAE and children’s faces including exposures up to 3 months before pregnancy. Previous studies show PAE before pregnancy is associated with other aspects of child developments, and the association is explained by maternal metabolic disorders such as impaired maternal glucose homeostasis and hepatic steatosis.^31–33^ The mechanism of the association with the face could be similar, but further investigations are needed to test this.

We compared the associations of PAE with facial morphology between the multi-ethnic group (Dutch, non-Dutch Western, Turkish and Moroccan) and the Western group (Dutch, non-Dutch Western), in which ethnicity was included as a covariate in the statistical analysis. The association we found were similar across ethnic groups. This suggests that the findings in the multi-ethnic group are not driven by the ethnicity, i.e., the linear regression model successfully adjusted for ethnicity as confounding bias.

### Strengthens and limitations

This study is a well-described population-based prospective cohort of multi-ethnic children. We have a large sample size with detailed PAE data that allows exposure classifications not available in many other studies. The deep-learning approaches exploited in this study represent the state-of-the-art method for high-dimensional 3D facial shape analysis.^21,34^ We integrated it with traditional linear and logistic regression models, to better interpret and validate the results in a conventional manner. Benefiting from these settings, the method used in this study is sensitive enough to detect association of mild alcohol consumption.

We also used derived endophenotypes for the prediction of PAE. Although the AUC increase with facial endophenotypes was rather small, the result still indicated that specific facial phenotypes provided additional and independent clues for association of PAE, which were confirmed by the odds ratio and *F*-tests **(Table S4** & **S5)**. Moreover, this shows that such a facial endophenotype approach could be valuable to study or diagnose other type of conditions.

Craniofacial development closely corresponds to brain development.^42^ Thus, classic facial features of FASD such as short palpebral fissure, smooth philtrum, and thin upper lip have been linked to brain abnormalities and cognitive outcome in FASD and have been used to diagnose children at risk of developing neurobehavioral deflicts.^9,37,38^ Low-moderate PAE has also shown adverse associations with children’s brain cognitive development, resulting in psychological and behavioral problems.^39–41^ Although the connection between these cognitive problems and facial phenotypes is still unknown, the endophenotypes we discovered in this study are potentially useful to assist in identifying children at risk of developing these cognitive problems, which should be further substantiated in future studies.

We had no data for alcohol consumption more than three months prior to pregnancy and thus do not know if maternal drinking had chronic effects. The self-reported questionnaire might not reflect the accuracy alcohol measurements because mothers may have denied their alcohol consumption.

## CONCLUSIONS

The results of this study suggest that low to moderate maternal alcohol consumption up to three months before and during the pregnancy is associated with the facial appearance of children. The association with facial morphology of the offspring was attenuated with increasing age. Future studies must show if facial morphology is related to child health and development. Our results imply that facial morphology, such as quantified by the approach we proposed here, can be used as a biomarker in further investigations. Furthermore, our study suggests that for women who are pregnant or want to become pregnant soon, should quit alcohol consumption several months before conception and completely during pregnancy to avoid adverse health outcomes in the offspring.

## Data Availability

private data from the Generation R study

## ACKNOWLEDGEMENTS

The Generation R Study is conducted by the Erasmus MC in close collaboration with the School of Law and Faculty of Social Sciences of the Erasmus University Rotterdam, the Municipal Health Service Rotterdam area, Rotterdam, the Rotterdam Homecare Foundation, Rotterdam and the Stichting Trombosedienst & Artsenlaboratorium Rijnmond (STAR-MDC), Rotterdam. We gratefully acknowledge the contribution of children and parents, general practitioners, hospitals, midwives, and pharmacies in Rotterdam.

## Contribution of Authorship

All authors have made significant contributions to this scientific work and approved the final version of the manuscript. XL was involved in the conception and design of the study, performed the data analyses and wrote the manuscript. GR supervised the data analyses, and co-wrote the manuscript. MK made contribution to result discussion, result interpretation and manuscript writing. HT made critical revisions on the manuscripts and provided extensive feedbacks. VWVJ, FR, SAK, WN and EW were involved in the conception and design of the study, reviewed the manuscript, and provided consultation regarding the analyses and interpretation of the data.

## Details of Ethics Approval

The study was approved by the Medical Ethics Committee (MEC) of the Erasmus Medical Center Rotterdam, The Netherlands (MEC 198.782/2001/31), and written informed consent was obtained from all participants themselves, or on that of their guardians/parents.

