## Supplementary for "Association between prenatal alcohol exposure and children’s facial shape. A prospective population-based cohort study"


| Content | Page number |
|---|---|
| Details about data pre-processing | 2 |
| Implementation details about 3D graph convolutional networks | 3 |
| Interpretation of facial endophenotypes | 4 |
| Details about the study population of different levels of prenatal alcohol exposure (PAE) | 7 |
| Details about dose-response assessment | 8 |
| Details about nominal significant endophenotypes | 9 |
| Detailed visualization results | 11 |
| Detailed PAE prediction results | 12 |

#### 1.Details about data pre-processing

1) **Data collection:** The 3D images were collected via the 3dMD cameras system (3dMD Corp). The raw data are triangle mesh with ~60,000 vertices and ~100,000 triangular faces.

2) **Data alignment I:** All raw 3D data were first aligned to the same position and orientation, where the subject's head and face pointed towards the y-axis and z-axis respectively. This was implemented by Matlab function 'Rigid ICP Registration'.[1]

3) **Landmarking, facial cropping and template setting:** The 3D coordinates of landmarks were calculated, and used to crop the facial regions from the raw data. Firstly, we did the cropping on 200 random subjects. Then a template was made by averaging these 200 samples.

4) **Down-sampling for the template:** The template had about 20,000 vertices and 40,000 triangular faces. We exploited the open-source ACVD[3] package to down-sample the template into 5,023 vertices and 9,851 triangular faces. The down-sampled template maintained the essential morphology while having a slight size.

5) **Landmark-guide dense correspondence:** In this step, we deformed the template by rigid and non-rigid registration algorithm,[2] in order to approximate the aligned images from step 2).

6) **Data alignment II:** We aligned all template-based results from step 5), and produced the final data set.

#### 2.Details about 3D graph convolutional networks

##### *Implementation details*

3D graph convolutional networks is an extension of 2D convolution networks. This extension allows deep learning networks to cope with 3D graph data. We used Gong's[4] framework in our study, and we kept his default configuration except for setting the latent size to be 200. We trained the networks on 9,017 data points of both 9-year-old and 13-year-old data, and stopped the training after 600 epochs. The training was based on self-supervised, and no labels were involved. After training, the parameters of the networks, together with the 200 latent endophenotypes of each facial shape, were saved as files.

##### *Reconstruction quality*

Higher reconstruction quality means better low-dimension representation of 3D input in the latent features. We computed a heatmap to display the reconstruction error. We also did experiments on different latent sizes, in order to understand how the latent size influenced the reconstruction quality. Then, we made a trade-off between reconstruction error and dimensional complexity and set the latent size as 200 in this study.

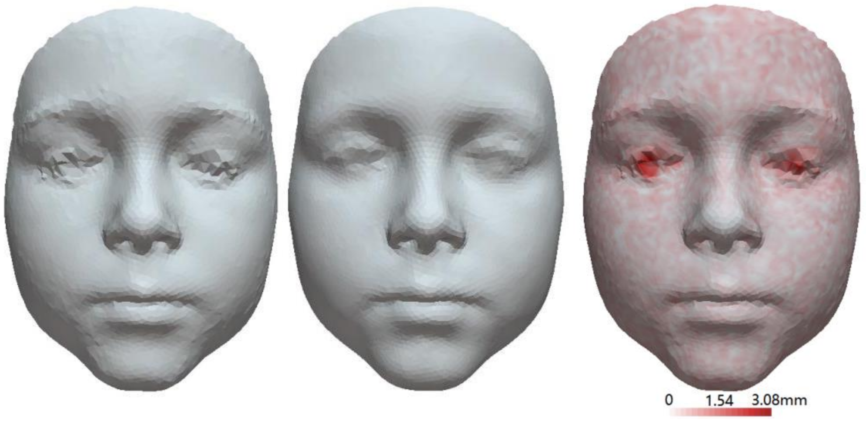

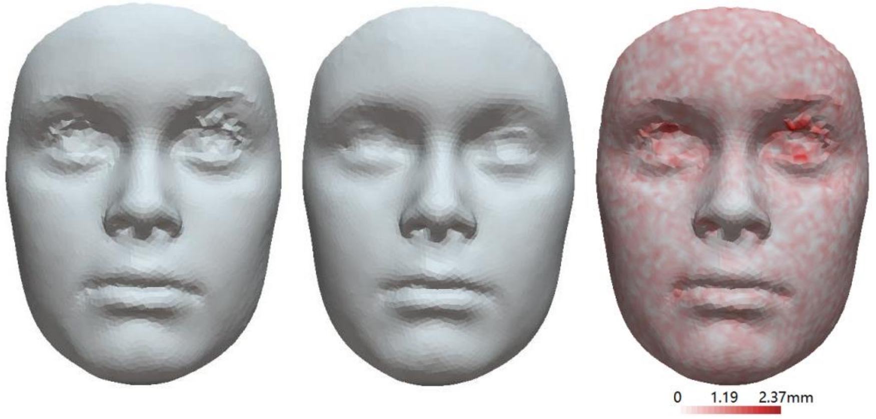


**Figure S1:** Reconstruction quality. left: input face; middle: reconstructed face; right: reconstruction error.

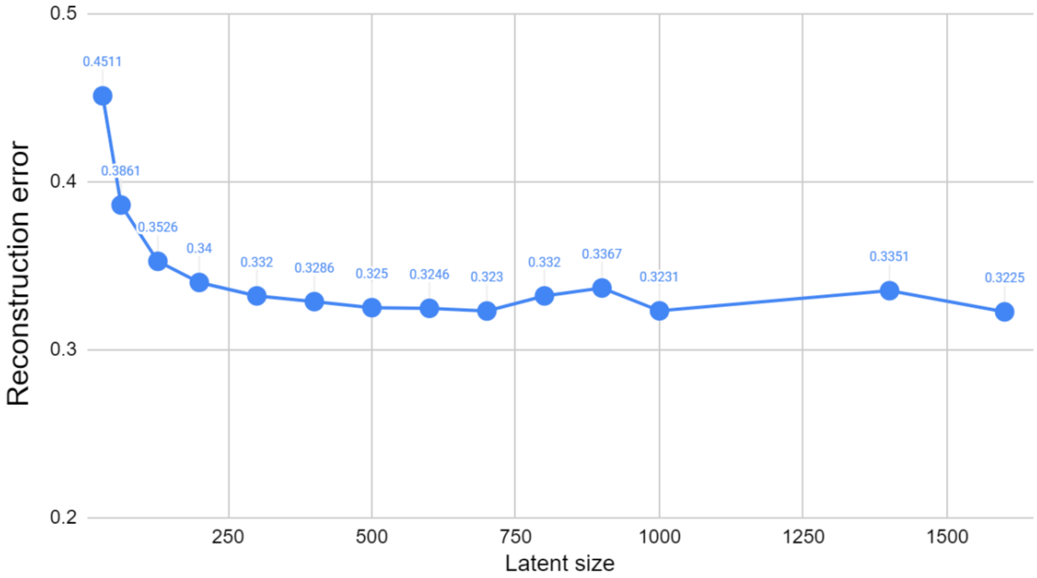


**Figure S2:** Reconstruction error of different latent sizes.

##### *Interpretation of 200 facial endophenotypes*

After training the networks, the 200 endophenotypes are generated by the Encoders, and each of them represents different structures on the 3D face. To better understand what each endophenotype is representing, in the decoding stage, we changed the value of one single endophenotype and observed the corresponding facial changes (**Figure S3a**). Furthermore, since each endophenotype may be corresponding to different amounts of facial changes, there is unfairness between each endophenotype. To address this, we defined a uniform value, $f(z)$, to measure the effect size of each endophenotype on the face:

$$f(z_i) = Sum\_Dist(\,Decode([\mu_0, \mu_1, \dots, \mu_i, \dots, \mu_{199}]), Decode([\mu_0, \mu_1, \dots, \mu_i + \sigma_i, \dots, \mu_{199}])\,) \qquad (S1)$$

where $\mu_i$ and $\sigma_i$ refer to the mean and standard deviation of $z_i(N = N_{sample})$ respectively, and $Sum\_Dist()$ is defined as the sum of Euclidean distance between all paired points from two 3D faces.

As shown in **Figure S3**, the interpretation is displayed by heatmaps, where red areas mean facial changes closer to the reference point, while blue areas mean changes farther to the reference point.

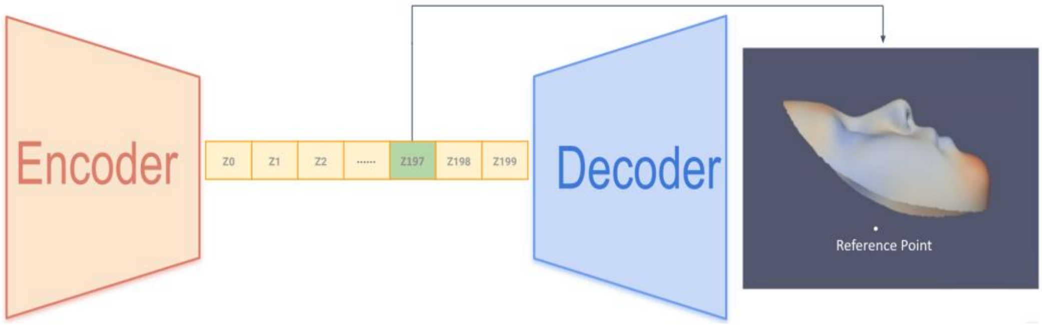


(a) Facial representation of one single endophenotype

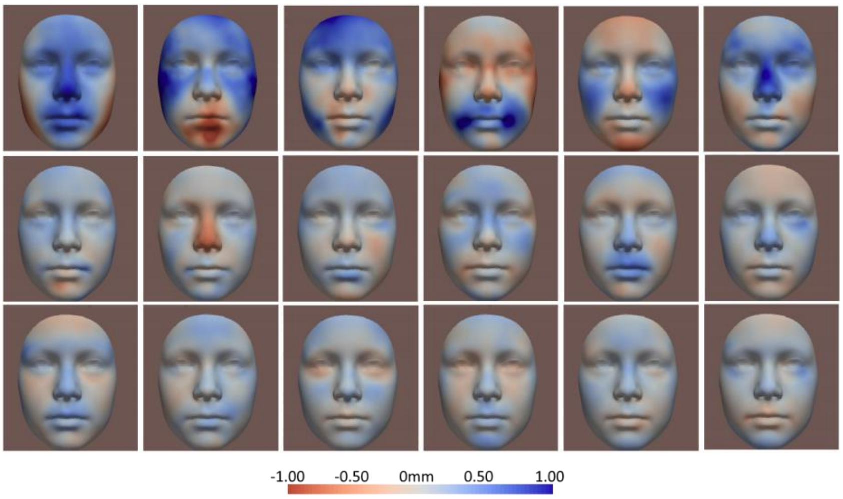


(b) Interpretation of each endophenotype

**Figure S3:** Interpretation of 200 endophenotypes. **a)** Facial changes corresponding to one endophenotype; **b**) Interpretation of each single endophenotype, sorted from major to minor facial changes based on the values of f(z).

##### ***Mapping selected endophenotypes back to the 3D face***

After selecting endophenotypes with statistical significance, we computed their corresponding effects on the 3D face. This process is similar with **Figure S3a**, but we computed the corresponding effects from multiple endophenotypes rather than single endophenotypes. As discussed, we performed Linear Regression on each endophenotypes $z_i$:

$$z_i = \beta_{i0} + \beta_{i1}x_{PAE} + \beta_{i2}x_{Materal\_age} + \beta_{i3}x_{Materal\_smoking} + \beta_{i4}x_{Children\_age} + \beta_{i5}x_{Children\_BMI} + \beta_{i6}x_{Chidlren\_gender} + \beta_{i7}x_{Children\_ethnicity} \quad i = 0,1,2 \ldots,199 \quad (S2)$$

**Equation S2** indicates that, the difference between non-exposed and exposed faces is corresponding to a change of $\beta_{i1}$ on $z_i$. Base on this, we combined all significant endophenotypes by setting $\beta_{i1}$ as their weights. As shown in **Figure S4**, in the decoding stage, we changed the values of all selected endophenotypes at the same time, and thus map these endophenotypes back to the 3D face:

$$\boldsymbol{F_{baseline}} = Decode([\mu_0, \mu_1, \mu_2, \dots, \mu_i, \dots, \mu_{199}]) \quad (S3)$$

$$\boldsymbol{F_{changed}} = Decode([\mu_0, \mu_1 + \beta_{11}, \mu_2 + \beta_{21}, \dots, \mu_i + \beta_{i1}, \dots, \mu_{199}]) \quad (S4)$$

$$Heatmap = PDist(\boldsymbol{F_{change}}, ref\_point) - PDist(\boldsymbol{F_{baseline}}, ref_{point}) \quad (S5)$$

where $\mu_i$ refers to the mean of $z_i$ ($N = N_{sample}$), $\beta_{11}$, $\beta_{21}$ and $\beta_{i1}$ are coefficients from the regression results, $\boldsymbol{F_{baseline}}$ is the average 3D face while $\boldsymbol{F_{change}}$ is the face affected by selected endophenotypes, $PDist()$ is defined as the point-wise Euclidean distance between each point on the 3D face and the reference point.

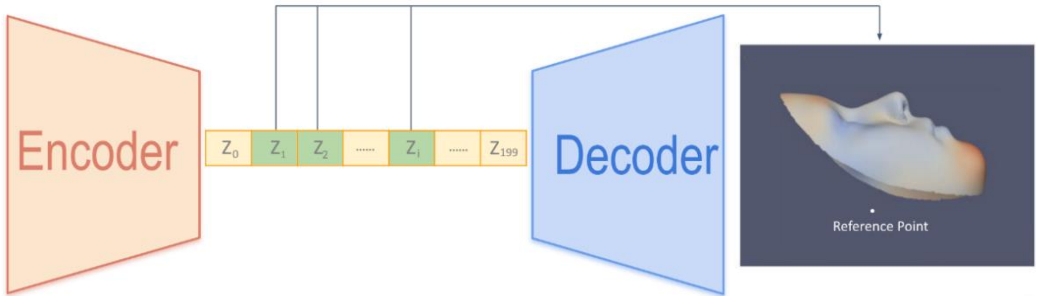


**Figure S4:** Combination of selected endophenotypes.

#### 3.Details about the study population of different levels of prenatal alcohol exposure (PAE)

**Table S1:** Details about alcohol consumption.
Level1: <1 drink per week;
Level2: 1–3 per week;
Level3: 4–6 per week;
Level4: 1 per day;
Level5: 2–3 per day;
Level6: >3 per day.
An average alcoholic drink contains about 12 g of alcohol.

| | 9-year-old | | 13-year-old | |
|---|---|---|---|---|
| **characteristic** | **Non-exposed (Control)** | **Any Alcohol** | **Non-exposed (Control)** | **Any Alcohol** |
| Alcohol exposure (%) | | | | |
| PAE Tier 1 | | | | |
| Drank alcohol before pregnancy only, level = 1 | 760 | 428 (60.6) | 519 | 349 (60.3) |
| Drank alcohol before pregnancy only, level = 2 | 760 | 158 (22.4) | 519 | 134 (23.1) |
| Drank alcohol before pregnancy only, level = 3 | 760 | 78 (11.0) | 519 | 56 (9.7) |
| Drank alcohol before pregnancy only, level = 4 | 760 | 28 (4.0) | 519 | 23 (4.0) |
| Drank alcohol before pregnancy only, level = 5 | 760 | 11 (1.6) | 519 | 15 (2.6) |
| Drank alcohol before pregnancy only, level = 6 | 760 | 3 (0.4) | 519 | 2 (0.3) |
| PAE Tier 2a | | | | |
| During pregnancy, any alcohol, level = 1 | 760 | 445(44.1) | 519 | 360(43.8) |
| During pregnancy, any alcohol, level = 2 | 760 | 354(35.1) | 519 | 286(34.8) |
| During pregnancy, any alcohol, level = 3 | 760 | 111(11.0) | 519 | 103(12.5) |
| During pregnancy, any alcohol, level = 4 | 760 | 46(4.6) | 519 | 27(3.3) |
| During pregnancy, any alcohol, level = 5 | 760 | 47(4.7) | 519 | 43(5.2) |
| During pregnancy, any alcohol, level = 6 | 760 | 5(0.5) | 519 | 3(0.4) |
| PAE Tier 2b | | | | |
| During pregnancy, any alcohol, level = 1 | 760 | 927(55.1) | 519 | 759 (55.0) |
| During pregnancy, any alcohol, level = 2 | 760 | 518(30.8) | 519 | 419 (30.4) |
| During pregnancy, any alcohol, level = 3 | 760 | 125(7.4) | 519 | 118 (8.6) |
| During pregnancy, any alcohol, level = 4 | 760 | 58(3.4) | 519 | 36 (2.6) |
| During pregnancy, any alcohol, level = 5 | 760 | 50(3.0) | 519 | 44 (3.2) |
| During pregnancy, any alcohol, level = 6 | 760 | 5(0.3) | 519 | 3 (0.2) |

### 4.Details about dose-response assessment

**Table S2:** FDR-significant endophenotypes of different levels of PAE, Tier 2b, multi-ethnic for 9-year-old children. Ne refers to the number of the exposed samples, while Nc refers to the number of the control samples.

**(a)** PAE level 1. Ne = 887, Nc = 760.

| endophenotypes Index | p-value | FDR-corrected p-value | coefficient | SE | Mean | Std | f(z) |
|---|---|---|---|---|---|---|---|
| 125 | 9.7e-05 | 0.019 | 0.016 | 0.0040 | 0.016 | 0.066 | 287.89 |
| 51 | 1.0e-04 | 0.010 | -0.029 | 0.0075 | -0.002 | 0.125 | 833.34 |
| 173 | 6.1e-04 | 0.041 | -0.011 | 0.0032 | -0.006 | 0.050 | 185.23 |

**(a)** PAE level 2-3. Ne = 643, Nc = 760.

| endophenotypes Index | p-value | FDR-corrected p-value | coefficient | SE | Mean | Std | f(z) |
|---|---|---|---|---|---|---|---|
| 139 | 8.1e-05 | 0.016 | -0.013 | 0.0034 | 0.007 | 0.048 | 189.49 |
| 51 | 1.2e-04 | 0.012 | -0.033 | 0.0085 | -0.002 | 0.125 | 833.34 |
| 36 | 6.1e-04 | 0.041 | 0.016 | 0.0045 | 0.010 | 0.064 | 269.78 |
| 29 | 9.4e-04 | 0.047 | 0.011 | 0.0034 | 0.008 | 0.052 | 197.95 |

**(a)** PAE level 4-6. Ne = 79, Nc = 760.

| endophenotypes Index | p-value | FDR-corrected p-value | coefficient | SE | Mean | Std | f(z) |
|---|---|---|---|---|---|---|---|
| 173 | 3.7e-05 | 0.0073 | -0.026 | 0.0063 | -0.006 | 0.050 | 185.23 |
| 157 | 2.5e-04 | 0.025 | -0.026 | 0.0070 | 0.003 | 0.054 | 220.76 |
| 36 | 8.7e-04 | 0.027 | 0.030 | 0.0084 | 0.010 | 0.064 | 269.78 |
| 29 | 8.7e-04 | 0.044 | 0.021 | 0.0063 | 0.008 | 0.052 | 197.95 |
| 69 | 1.1e-03 | 0.046 | 0.035 | 0.011 | -0.017 | 0.080 | 485.34 |

#### 5.Details about nominal significant endophenotypes

**Table S3:** Details about nominal significant endophenotypes, Tier 2b, PAE level > 1, multi-ethnic.
Ne refers to the number of the exposed samples, while Nc refers to the number of the control samples.
As defined in Equation S1, f(z) is the effect size of the endophenotype on the facial shape.
Index with * was still significant after adjusted by FDR.

**(a)** 9-year-old children. Ne = 756, Nc = 760

| endophenotypes Index | p-value | FDR-corrected p-value | coefficient | SE | Mean | Std | f(z) |
|---|---|---|---|---|---|---|---|
| 36* | 7.1e-05 | 0.014 | 0.017 | 0.0044 | 0.010 | 0.064 | 269.78 |
| 139* | 9.3e-05 | 0.009 | -0.013 | 0.0033 | 0.007 | 0.048 | 189.49 |
| 29* | 0.0002 | 0.013 | 0.013 | 0.0034 | 0.008 | 0.052 | 197.95 |
| 51* | 0.0002 | 0.012 | -0.030 | 0.0083 | -0.002 | 0.125 | 833.34 |
| 69* | 0.0006 | 0.023 | 0.019 | 0.0055 | -0.017 | 0.080 | 485.34 |
| 173* | 0.0009 | 0.030 | -0.011 | 0.0033 | -0.006 | 0.050 | 185.23 |
| 87* | 0.001 | 0.036 | -0.010 | 0.0031 | 0.000 | 0.044 | 155.85 |
| 57* | 0.002 | 0.048 | 0.022 | 0.0070 | -0.013 | 0.101 | 648.42 |
| 125 | 0.005 | 0.101 | 0.013 | 0.0045 | 0.016 | 0.066 | 287.89 |
| 12 | 0.005 | 0.092 | -0.037 | 0.0129 | 0.070 | 0.208 | 1852.95 |
| 178 | 0.006 | 0.102 | 0.010 | 0.0036 | 0.000 | 0.054 | 209.58 |
| 58 | 0.006 | 0.107 | 0.011 | 0.0041 | 0.007 | 0.059 | 255.12 |
| 174 | 0.007 | 0.106 | 0.016 | 0.0058 | 0.040 | 0.092 | 493.61 |
| 163 | 0.009 | 0.132 | 0.010 | 0.0038 | -0.002 | 0.054 | 214.16 |
| 97 | 0.010 | 0.135 | -0.015 | 0.0058 | 0.012 | 0.082 | 419.65 |
| 55 | 0.011 | 0.135 | -0.012 | 0.0048 | 0.005 | 0.068 | 285.65 |
| 75 | 0.012 | 0.141 | 0.008 | 0.0033 | -0.006 | 0.048 | 181.64 |
| 121 | 0.018 | 0.198 | -0.009 | 0.0037 | -0.007 | 0.057 | 227.44 |
| 83 | 0.018 | 0.188 | -0.009 | 0.0036 | 0.006 | 0.053 | 190.46 |
| 39 | 0.018 | 0.180 | 0.012 | 0.0049 | -0.002 | 0.072 | 345.68 |
| 54 | 0.024 | 0.224 | 0.008 | 0.0036 | -0.008 | 0.053 | 221.60 |
| 53 | 0.026 | 0.239 | 0.007 | 0.0032 | 0.019 | 0.047 | 177.26 |
| 19 | 0.035 | 0.305 | 0.012 | 0.0055 | -0.012 | 0.086 | 484.90 |
| 47 | 0.040 | 0.330 | -0.007 | 0.0032 | 0.004 | 0.047 | 178.17 |
| 160 | 0.042 | 0.332 | -0.008 | 0.0037 | -0.008 | 0.053 | 203.75 |
| 185 | 0.042 | 0.320 | -0.007 | 0.0033 | 0.002 | 0.049 | 172.72 |
| 7 | 0.042 | 0.314 | -0.007 | 0.0036 | 0.005 | 0.053 | 214.33 |
| 128 | 0.046 | 0.330 | -0.006 | 0.0031 | -0.003 | 0.046 | 148.50 |

**(b)** 13-year-old children. Ne = 620, Nc = 519

| endophenotypes Index | p-value | FDR-corrected p-value | coefficient | SE | Mean | Std | f(z) |
|---|---|---|---|---|---|---|---|
| 74 | 0.0004 | 0.071 | 0.018 | 0.0050 | -0.011 | 0.066 | 269.42 |
| 12 | 0.0006 | 0.064 | -0.061 | 0.0179 | -0.028 | 0.245 | 2185.73 |
| 45 | 0.003 | 0.182 | 0.013 | 0.0045 | 0.009 | 0.057 | 213.14 |

| | | | | | | | |
|---|---|---|---|---|---|---|---|
| 193 | 0.005 | 0.239 | 0.018 | 0.0064 | -0.018 | 0.084 | 414.69 |
| 85 | 0.007 | 0.272 | -0.011 | 0.0040 | 0.001 | 0.052 | 176.85 |
| 164 | 0.007 | 0.247 | -0.012 | 0.0043 | 0.001 | 0.054 | 204.62 |
| 14 | 0.011 | 0.303 | 0.031 | 0.0122 | 0.013 | 0.158 | 1126.57 |
| 46 | 0.012 | 0.289 | 0.009 | 0.0037 | 0.008 | 0.048 | 176.55 |
| 90 | 0.012 | 0.258 | -0.014 | 0.0055 | 0.013 | 0.072 | 343.16 |
| 172 | 0.012 | 0.237 | -0.010 | 0.0038 | 0.002 | 0.050 | 170.99 |
| 26 | 0.012 | 0.218 | -0.011 | 0.0043 | -0.001 | 0.056 | 226.21 |
| 5 | 0.013 | 0.219 | -0.011 | 0.0044 | 0.015 | 0.055 | 248.72 |
| 196 | 0.015 | 0.234 | -0.021 | 0.0088 | 0.008 | 0.116 | 674.34 |
| 16 | 0.017 | 0.242 | -0.013 | 0.0053 | -0.007 | 0.067 | 308.31 |
| 66 | 0.017 | 0.227 | 0.009 | 0.0036 | 0.003 | 0.049 | 180.30 |
| 51 | 0.024 | 0.296 | -0.022 | 0.0098 | 0.034 | 0.135 | 898.73 |
| 197 | 0.025 | 0.290 | -0.008 | 0.0037 | -0.009 | 0.047 | 168.62 |
| 7 | 0.027 | 0.296 | -0.009 | 0.0040 | -0.003 | 0.051 | 207.37 |
| 87 | 0.034 | 0.357 | -0.008 | 0.0039 | 0.001 | 0.050 | 176.29 |
| 75 | 0.035 | 0.351 | 0.008 | 0.0036 | -0.006 | 0.047 | 179.60 |
| 109 | 0.043 | 0.405 | -0.008 | 0.0041 | -0.006 | 0.053 | 223.21 |
| 80 | 0.043 | 0.392 | 0.009 | 0.0043 | -0.004 | 0.054 | 211.15 |
| 96 | 0.045 | 0.391 | -0.011 | 0.0054 | 0.018 | 0.072 | 325.94 |
| 47 | 0.049 | 0.407 | -0.007 | 0.0037 | -0.002 | 0.048 | 179.54 |

**(c)** ‘growth’. Ne = 460, Nc = 377.

| endophenotypes Index | p-value | FDR-corrected p-value | coefficient | SE | Mean | Std | f(z) |
|---|---|---|---|---|---|---|---|
| 12 | 0.017 | 0.236 | -0.040 | 0.0168 | 0.026 | 0.231 | 2053.41 |
| 16 | 0.008 | 0.761 | -0.014 | 0.0052 | 0.005 | 0.065 | 299.80 |
| 19 | 0.022 | 0.850 | -0.018 | 0.0079 | -0.010 | 0.094 | 531.21 |
| 36 | 0.013 | 0.702 | -0.015 | 0.0061 | 0.012 | 0.063 | 267.30 |
| 43 | 0.028 | 0.591 | 0.009 | 0.0042 | 0.002 | 0.045 | 162.83 |
| 53 | 0.001 | 0.544 | -0.015 | 0.0046 | 0.012 | 0.047 | 175.87 |
| 66 | 0.028 | 0.472 | 0.010 | 0.0046 | 0.001 | 0.048 | 177.34 |
| 74 | 0.037 | 0.430 | 0.011 | 0.0054 | -0.005 | 0.064 | 261.61 |
| 81 | 0.023 | 0.479 | -0.021 | 0.0093 | -0.004 | 0.094 | 527.73 |
| 93 | 0.036 | 0.463 | 0.010 | 0.0050 | -0.002 | 0.050 | 187.42 |
| 97 | 0.015 | 0.505 | 0.018 | 0.0072 | 0.017 | 0.085 | 431.61 |
| 139 | 0.014 | 0.469 | 0.011 | 0.0044 | 0.005 | 0.049 | 194.45 |
| 156 | 0.044 | 0.526 | 0.010 | 0.0048 | 0.000 | 0.049 | 185.81 |
| 164 | 0.017 | 0.510 | -0.012 | 0.0050 | -0.002 | 0.054 | 203.93 |
| 172 | 0.016 | 0.491 | -0.011 | 0.0046 | 0.003 | 0.049 | 169.76 |
| 183 | 0.034 | 0.545 | 0.013 | 0.0062 | 0.004 | 0.062 | 270.37 |

### 6. Detailed visualization results

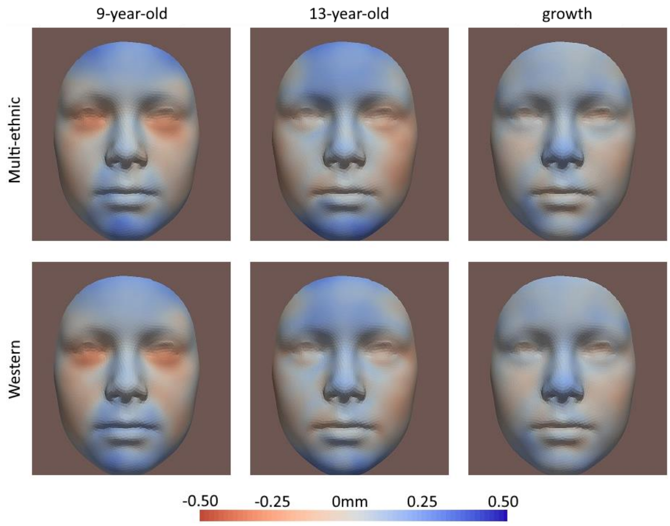


**Figure S5:** Nominal significant results (p-value<0.05). Tier 2b, PAE level > 1.

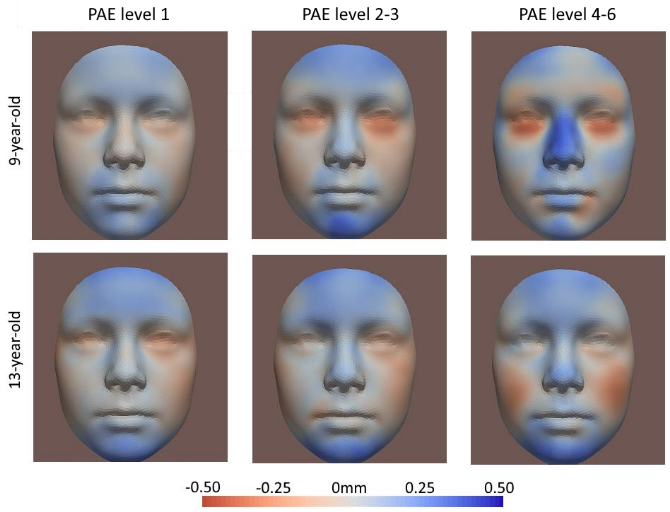


**Figure S6:** Nominal significant results of different levels of PAE. Tier 2b for multi-ethnic group.

#### 7. Detailed PAE prediction results

**Table S4:** Sorted odds ratio of top 8 endophenotypes.
Ne refers to the number of the exposed samples, while Nc refers to the number of the control samples.

| **9-year-old** (Ne = 670, Nc = 329) | | | **13-year-old** (Ne = 543, Nc = 236) | | |
|---|---|---|---|---|---|
| **Endophenotype Index** | **odds ratio** | **p-value** | **Endophenotype Index** | **odds ratio** | **p-value** |
| 36 | 1.25 | 0.0077 | 14 | 1.36 | 0.0039 |
| 57 | 1.16 | 0.065 | 74 | 1.33 | 0.0060 |
| 39 | 1.15 | 0.11 | 45 | 1.28 | 0.013 |
| 125 | 1.15 | 0.10 | 96 | 0.88 | 0.18 |
| 69 | 1.13 | 0.16 | 12 | 0.86 | 0.15 |
| 139 | 0.87 | 0.10 | 85 | 0.86 | 0.11 |
| 83 | 0.82 | 0.016 | 87 | 0.81 | 0.027 |
| 87 | 0.76 | 0.00083 | 164 | 0.76 | 0.0069 |

**Table S5:** Sorted *F*-test results of top 8 endophenotypes.

| **9-year-old** (Ne = 670, Nc = 329) | | | **13-year-old** (Ne = 543, Nc = 236) | | |
|---|---|---|---|---|---|
| **Endophenotype Index** | **p-value** | **F-value** | **Endophenotype Index** | **p-value** | **F-value** |
| 39 | 7.6e-6 | 20.3 | 12 | 3.5e-7 | 26.4 |
| 139 | 9.7e-5 | 15.3 | 164 | 1.8e-4 | 14.2 |
| 87 | 1.5e-4 | 14.5 | 74 | 1.8e-4 | 14.1 |
| 36 | 3.4e-4 | 12.9 | 85 | 3.8e-4 | 12.7 |
| 57 | 5.3e-4 | 12.1 | 96 | 9.6e-3 | 6.7 |
| 125 | 2.1e-3 | 9.5 | 14 | 1.6e-2 | 5.9 |
| 69 | 1.1e-2 | 6.4 | 87 | 1.6e-2 | 5.8 |
| 83 | 0.077 | 3.1 | 45 | 0.29 | 1.1 |

**1.** Manu (2020). Rigid ICP registration (https://www.mathworks.com/matlabcentral/fileexchange/40888-rigid-icp-registration), *MATLAB Central File Exchange*. Retrieved October 14, 2020.

**2.** Amberg B, Romdhani S and Vetter T. Optimal Step Nonrigid ICP Algorithms for Surface Registration, *IEEE Conference on Computer Vision and Pattern Recognition.* Minneapolis, MN, 2007, pp. 1-8, doi: 10.1109/CVPR.2007.383165.

**3.** Valette S, Chassery JM and Prost R. Generic remeshing of 3D triangular meshes with metric-dependent discrete Voronoi Diagrams, *IEEE Transactions on Visualization and Computer Graphics*. 2008; 14(2):369-381

**4.** Gong S, Chen L, Bronstein M and Zafeiriou S. SpiralNet++: A Fast and Highly Efficient Mesh Convolution Operator, *IEEE/CVF International Conference on Computer Vision Workshop (ICCVW)*, Seoul, Korea (South), 2019, pp. 4141-4148, doi: 10.1109/ICCVW.2019.00509
